# Efficacy of tDCS and EEG Neurofeedback, individually and combined, on Neuropathic Pain following spinal cord injury: Protocol for a Randomised Controlled Trial

**DOI:** 10.64898/2026.03.11.26347999

**Authors:** Nahian S Chowdhury, Negin Hesam-Shariati, Yann Quide, Pauline Zahara, Robert Herbert, Sebastian Restrepo, Kevin Chen, Angus Mcyntire, Toby Newton John, James W Middleton, Ashley Craig, Mark P Jensen, Jane Butler, Nancy Briggs, James McAuley, Sylvia M Gustin

## Abstract

Neuropathic pain (NP) affects approximately 60% of individuals with spinal cord injury (SCI). Existing pharmacological treatments provide only modest relief and are often limited by adverse effects, while non-pharmacological options show small effects at best. As such, there remains a need for accessible, mechanism-informed treatments for SCI-NP. This protocol describes a trial evaluating two promising home-based neuromodulatory interventions for SCI-NP - electroencephalography neurofeedback (EEG-NF) and transcranial direct current stimulation (tDCS) - tested both independently and when applied in combination. We will employ a partially double-blinded (i.e. 1 treatment blinded, the other not), 2×2 factorial randomised controlled trial. Adults with chronic SCI-NP (N=192) will be randomised to: (1) EEG-NF + active tDCS, (2) EEG-NF + sham tDCS, (3) active tDCS alone, or (4) sham tDCS alone, in addition to treatment as usual. Participants will complete 20 home-based sessions over 5 weeks. The primary outcome is change in overall pain severity with the primary endpoint being 6 weeks post-randomisation, with secondary endpoints at 16, 26 and 52 weeks post-randomisation. Secondary outcomes (worst pain intensity, pain interference, sleep, depressive symptoms, health-related quality of life) will be assessed at 6 weeks, 16 weeks, 26 weeks and 52 weeks post-randomisation. This will be the first large-scale trial of home-based EEG-NF and tDCS for SCI-NP. If found to be effective, these scalable interventions could be integrated into routine care and inform further optimisation of neuromodulation strategies for managing SCI-NP.

## Introduction

Neuropathic pain (NP) affects approximately 60% of individuals with spinal cord injury (SCI) and is often rated by these individuals as more disabling than loss of mobility^1^. It typically occurs at or below the injury level, and is described as burning, shock-like, tingling, stabbing sharp and persistent, and substantially impairs mood, sleep and social functioning^2^. Moreover, SCI-NP is often resistant to treatment and tends to worsen over time^3^. Current evidence-based pharmacological treatments for central neuropathic pain (tricyclic antidepressants, SNRIs, gabapentin and pregabalin) provide only modest pain relief on average and are frequently limited by adverse effects; opioids are generally discouraged, with tramadol reserved for short episodes of severe pain^4^. Non-pharmacological approaches, such as cognitive behavioural therapy, physical therapies, acupuncture, and therapeutic hypnosis generally show at best small effects, and, for central neuropathic pain specifically, there exists insufficient evidence^2–6^. The lack of effective treatments highlights the need for new treatments to address SCI-NP and improve patients’ pain outcomes. Here we present a protocol for a trial aiming to evaluate two promising home-based neuromodulation treatments for SCI-NP. Home-based treatments are gaining traction due to their potential to accommodate mobility limitations in individuals with SCI-NP.

### Electroencephalography (EEG) Neurofeedback (NF)

The development and maintenance of chronic neuropathic pain following SCI is hypothesized to be associated with thalamocortical dysrhythmia^7^. This problem can be detected using surface electroencephalography (EEG)^8^. In SCI-NP, this dysrhythmia has been characterised by increased resting state theta (4–7 Hz) and high beta (20–30 Hz) power and reduced sensorimotor rhythm (10–15 Hz) power compared with healthy controls^8–10^. EEG-NF trains individuals to self-regulate these rhythms.^11,12^ EEG is recorded (e.g., over sensorimotor cortex), relevant frequency bands are extracted in real time, and their power is translated into game-like or audio-visual feedback that reinforces desired brain activity^13–16^. This may enable individuals with SCI-NP to normalise dysrhythmic activity and reduce pain^8,17^. EEG-NF has shown promising effects for SCI-NP with a favourable adverse-effect profile compared to pharmacological therapies^8,12^. However, large, well-controlled randomised controlled trials (RCTs) in SCI-NP are lacking. A recent RCT in 116 individuals with chronic pain compared active EEG-NF to a sham “yoked feedback” condition where participants viewed another individual’s feedback; 44–46% of participants in both groups achieved moderate pain relief, with no between-group difference^18^. However, as the authors note, the sham condition likely acted as an active intervention, because ∼25% of the replayed feedback still coincided with desirable EEG patterns. Consistent with broader critiques of sham neurofeedback^19^, such designs may underestimate true treatment effects. To minimise this issue, the current trial evaluates EEG-NF in addition to treatment as usual, rather than against an active sham.

### Transcranial Direct Current Stimulation (tDCS)

Chronic neuropathic pain is often associated with an increase in activity in central pain pathways (central sensitisation^20^) motivating treatments that directly target the central nervous system. tDCS involves delivery of an electrical current through scalp electrodes, resulting in modulation of cortical excitability^21^, and activation of descending anti-nociceptive pathways^22^ with relatively few side effects compared with pharmacological or surgical options. In a 2018 Cochrane review of RCTs for chronic pain^23^ and 2025 Lancet review of RCTs for neuropathic pain^24^, while tDCS was found to elicit small to moderate reductions in pain intensity, the certainty of evidence was rated very low. As discussed in our recent perspective piece^25^, two factors for this inconclusive evidence are (1) underpowered sample sizes across trials and (2) a strong reliance on a generic tDCS montage (placement of electrodes) involving the anode placed on left M1 and cathode placed on the right supraorbital (SO) region^26^, rather than tailoring the montage towards the mechanisms underlying pain. Because SCI-NP is commonly dominated by bilateral and lower extremity pain, a montage that preferentially modulates the bilateral lower extremity motor areas in M1 may be a more appropriate mechanistic target^27^. Thus, in addition to adequate statistical power, the present trial will, for the first time, test a novel montage for SCI-NP with the anode positioned over the vertex and the return electrode on the shoulder. Modelling suggests that this configuration engages bilateral lower-extremity motor cortical representations, while the extracephalic cathode may reduce confounding cortical effects associated with a cephalic return electrode.

### Combined EEG-NF and tDCS

The combination of EEG-NF and tDCS may also yield therapeutic benefits. tDCS has been hypothesised to “prime” cortical networks by increasing cortical excitability and promoting plasticity, thereby enhancing the effects of concurrent therapies^28^. In SCI-NP, a study of 39 participants found that tDCS augmented the pain-relieving effects of a visual illusion gait intervention compared with either treatment alone^29^. A larger RCT of 130 participants reported greater pain reduction when tDCS was combined with visual illusions when compared to a control group, although it did not compare the combination to sham tDCS (i.e. placebo)^30^. These findings suggest that pairing tDCS with an EEG-NF may also yield therapeutic benefits, however no large-scale trial has yet tested this in SCI-NP.

### Aims of the Current Trial

The primary aim of the study described herein is to conduct an RCT that addresses the methodological limitations of prior research, described previously. Specifically, we plan to conduct a 2×2 factorial RCT that will evaluate the independent and combined efficacy of home-based EEG-NF and tDCS for SCI-NP. This will be the first large-scale trial of these home-based interventions in SCI-NP and will provide critical evidence to guide future clinical implementation.

## Methods

### Trial Design

The trial will use a partially double-blinded, 2 × 2 factorial randomised controlled design. Participants will be allocated to one of the four groups shown in Table 1.

**Table 1.** Group Allocation.

|  | Receives active tDCS | Receives sham tDCS |
| --- | --- | --- |
| Receives EEG NF | Group 1: EEG-NF + Active tDCS | Group 2: EEG-NF + Sham tDCS |
| Does not Receive EEG -NF | Group 3: Active tDCS | Group 4: Sham tDCS |

Participants in all groups will continue to receive their usual treatments and care throughout the study, as clinically indicated. This may include, but is not limited to, pharmacological, psychological and rehabilitative treatments. The interventions tested in this trial will be provided in addition to, not instead of, their current treatment

The trial has been prospectively registered with the Australian and New Zealand Clinical Trials Registry (ACTRN12625000737437p) and this protocol adheres to the Standard Protocol Items: Recommendations for Interventional Trials Guidelines^31^. Findings from this study will be reported according to the Consolidated Standards of Reporting Trials statement^32^. Figure 1 shows the CONSORT flow diagram.

**Figure 1.**
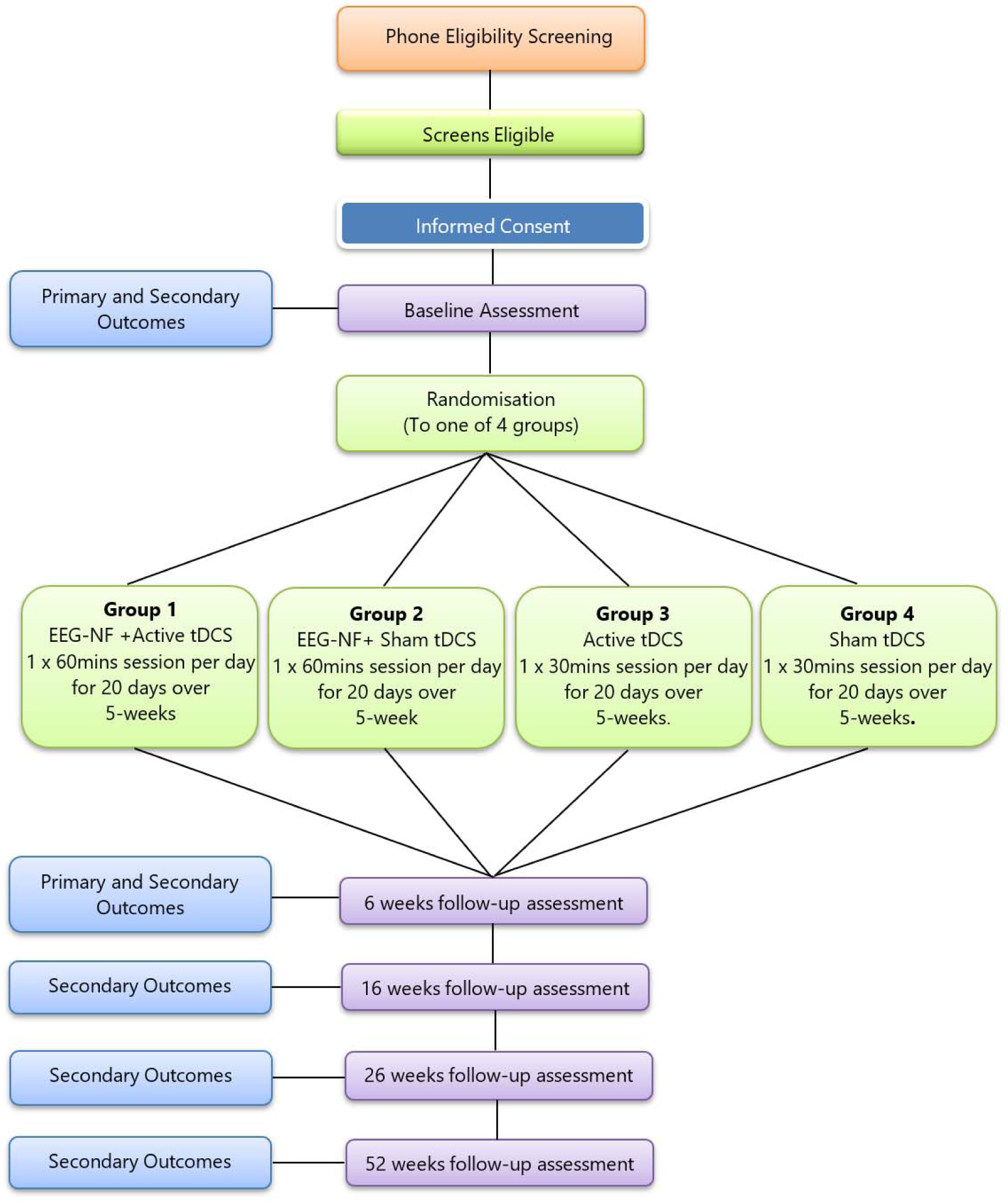
CONSORT flow diagram.

### Objectives

#### Primary Objectives

1. To test the causal effect of EEG-NF compared to no EEG-NF on pain severity at 6 weeks post-randomisation (primary endpoint), in individuals not given active tDCS.
2. To test the causal effect of active tDCS compared to sham tDCS on pain severity at 6 weeks post-randomisation, in individuals not given EEG-NF.
3. To test the causal effect of combined EEG-NF and active tDCS compared to sham tDCS on pain severity at 6 weeks post-randomisation.

#### Secondary Objectives

1. To describe, in individuals not given active tDCS, the possible causal effect of EEG**-**NF compared to no-EEG-NF on:

a. Pain severity at 16, 26, and 52 weeks post-randomisation.
b. Worst pain severity, pain interference, sleep problems, depressive symptoms and health-related quality of life at 6, 16, 26, and 52 weeks post-randomisation.
2. To describe, in individuals not given EEG-NF, the possible causal effect of active tDCS compared to sham tDCS on:

a. Pain severity at 16, 26, and 52 weeks post-randomisation.
b. Worst pain severity, pain interference, sleep problems, depressive symptoms and health-related quality of life at 6, 16, 26, and 52 weeks post-randomisation.
3. To describe the possible causal effect of combined EEG-NF and active tDCS compared to sham tDCS, on:

a. Pain severity at 16, 26, and 52 weeks post-randomisation.
b. Worst pain severity, pain interference, sleep problems, depressive symptoms and health-related quality of life at 6, 16, 26, and 52 weeks post-randomisation.

### Sample Size

The sample size calculation was conducted by Dr Robert Herbert, an independent biostatistician. The sample size required to provide nominal power for null hypothesis tests corresponding to each of the trial’s primary objectives was determined analytically and using Monte Carlo simulations (5,000 simulated trials). The trial is powered to detect a 1-point difference on the primary outcome (pain severity), consistent with the OMERACT-recommended minimal clinically important difference (MCID) of 1 point on a 0–10 scale^33^. The SD (1.9) was taken from a previous SCI neuropathic pain trial (HC210932, VRWalk^34^) and conservatively rounded to 2.0. The baseline–post correlation of the measure of pain severity in that trial was 0.76; here, correlations between adjacent timepoints were assumed to be 0.75, decaying by 0.05 for each increasingly distant timepoint. Baseline (pre-treatment) pain severity score was included as a covariate. The nominal Type I error rate (two-sided) was set at 0.05, and allowance was made for a worst-case loss to follow-up of 20%. These calculations indicated that a total sample size of 192 participants (48 per group) would provide 90% power for each of the three primary objectives (main effect of tDCS vs sham tDCS; main effect of EEG neurofeedback vs control; and their combined effect).

### Study setting and eligibility

Participants will complete the intervention at home anywhere in Australia; all equipment and procedure manuals will be mailed to their address. The study will recruit adults (≥18 years) with a complete or incomplete cervical, thoracic or lumbar spinal cord injury ≥6 months post-injury, with ongoing neuropathic pain at or below the injury level for >3 months and average pain ≥4/10 over the past week. Eligibility requires endorsement of ≥2 items on the 4-item Screening for SCI-NP Instrument (SCIPI)^35^, independent breathing, ability to read and understand English, and capacity to complete the intervention (self-administering tDCS/EEG-NF or with caregiver assistance), including one session per day for 20 days over 5 weeks. Exclusion criteria include contraindications to brain stimulation (e.g., cardiac pacemaker, cochlear implant, deep brain stimulator, metal in the head, history of serious head injury or stroke, personal or first-degree family history of seizures, pregnancy or lactation), neurological or psychiatric disorders with cognitive impairment (e.g., dementia, schizophrenia), and residence outside Australia.

### Recruitment and consent

Participants will be recruited using existing databases at Neuroscience Research Australia, NeuroRecovery Research Hub, UNSW, the George Institute’s Join Us register, as well as using community flyers, social media, and national SCI/NP organisations and support groups. Prospective participants will receive a Participant Information Statement, have time to consider participation, and provide electronic consent via REDCap with support from a research assistant trained in Good Clinical Practice. They may optionally provide consent for the release of Medicare Benefits Schedule (MBS) and Pharmaceutical Benefits Scheme (PBS) data from Services Australia for health economic analyses.

### Randomisation

Following eligibility screening, informed consent, and baseline questionnaires, participants will be randomised to one of four groups (Groups 1–4) in a 1:1:1:1 ratio. The randomisation sequence will be in permuted blocks of size 4, 8 or 12. An independent statistician, not involved in recruitment or assessment, will use a random number generator to create a schedule assigning each participant ID to a group. After consent and baseline assessment, allocations will be revealed (on REDCap) only to a team member not involved in recruitment or assessment, who will then mail the relevant equipment and manuals to participants’ home addresses. The first intervention session will commence 1 week later to allow for delivery. The allocation schedule will remain inaccessible to other trial staff until study completion to ensure allocation concealment.

### Blinding

Participants and assessors will be blinded to tDCS allocation; however, blinding will not be feasible for EEG-NF. An investigator independent of outcome assessment will pre-program the device to deliver active or sham tDCS. Sham involves a 20s ramp-up to 2 mA, a 17 s ramp-down to 0 mA, and brief 0.1 mA pulses every 5 minutes to monitor impedance, mimicking active sensations without ongoing current. Participants in all groups will be informed they may experience mild scalp sensations (e.g. tingling/itching) that may diminish over time. Participants will not be told their allocation and will be debriefed at study completion. Blinding integrity will be assessed via end-of-trial questionnaires in which participants guess their tDCS allocation and rate confidence, and assessors guess group allocation. All analyses will use de-identified, coded data, and the allocation schedule will remain inaccessible to other staff until trial completion to preserve concealment.

### Overview of Intervention

All participants will complete 20 home-based sessions over five weeks (four sessions per week), commencing no earlier than one-week post-randomisation. Groups 1 and 2 will complete 60-minute sessions combining tDCS and EEG-NF; Groups 3 and 4 will complete 30-minute tDCS-only sessions. For all groups, a study tablet will guide each session with step-by-step setup instructions, and for Groups 1 and 2, serve as the EEG-NF interface. Each session will include 10 minutes of tDCS setup followed by 20 minutes of stimulation. Groups 3 and 4 then conclude, whereas Groups 1 and 2 complete an additional 10-minute EEG-NF setup and 20 minutes of neurofeedback. To verify adherence and montage accuracy, the tablet will prompt all participants to upload a photo of their tDCS and EEG-NF setup before the intervention. Images will be uploaded in real time to a secure AWS Cloud dashboard for monitoring and quality assurance.

### tDCS Intervention

#### Overview

Active tDCS or Sham will be delivered at 2mA for 20 min via a Sooma stimulator (ARTG ID 278349, Sooma, Finland) using 5 cm circular rubber electrodes with saline-soaked sponge pads. Stimulation will commence only once electrode impedance is sufficiently low. If impedance is too high, the tablet interface will instruct participants to apply additional saline.

#### tDCS Montage

Existing RCTs for SCI-NP have placed the anode over C3 (hand motor cortex) and the cathode over the right supraorbital region^23^. However, systematic reviews have reported inconclusive evidence for neuropathic pain using this montage^24,36^, possibly because it does not specifically target neural mechanisms and body regions most relevant to SCI-NP. Converging evidence indicates that cortical excitability is a key mechanistic target for neuromodulation. Low corticomotor excitability has been proposed as a biomarker for chronic pain development^37^, and individuals with SCI-NP exhibit reduced motor cortex excitability compared with those without NP^38^. Furthermore, high-frequency rTMS, which increases cortical excitability^39^, has been shown to produce analgesic effects in neuropathic pain^40^. Because SCI-NP is commonly dominated by bilateral and lower-extremity pain, an optimal montage may be one that preferentially modulates the bilateral and lower-extremity motor cortical representations.

A review of tDCS montages^41^ highlighted work by Kasi et al.^42^ , showing that a Cz–Inion montage significantly increased motor-evoked potential amplitudes in bilateral tibialis anterior. Electrical field modelling further demonstrated that Cz–Inion tDCS aligns current flow parallel to pyramidal tract fibres projecting to the lower extremities, enhancing stimulation efficiency ^43^ and subsequent work in 31 participants confirmed more pronounced reductions in lower limb motor thresholds with Cz–Inion than with C3–C4 or C1–C2 montages. These data support Cz–Inion as a promising approach for increasing excitability in lower extremity representations.

However, montage selection must also avoid unwanted inhibitory effects at the cathode, which may attenuate or reverse anodal benefits^44^. Placing the cathode at the Inion delivers cathodal (inhibitory) stimulation over cerebellar regions, which may be suboptimal for pain modulation^45^. A recent synthesis of brain stimulation targets for chronic pain, combining a meta-analysis of 92 neuroimaging studies with resting-state fMRI in 90 patients, identified the cerebellum as a promising site for excitatory neuromodulation. Consistent with this, anodal cerebellar tDCS has reduced pain intensity during experimental pain in healthy volunteers and in individuals with peripheral neuropathic pain ^46,47^. Cathodal cerebellar stimulation, as in Cz–Inion, could therefore undermine analgesic aims.

To address this, extra-cephalic montages placing the return electrode on the shoulder have been proposed. Modelling studies indicate that extra-cephalic placements can generate more focused current under the anode and allow greater depth of penetration to reach deeper structures. Experimental work applying anodal tDCS over the vertex with a shoulder return electrode has shown significant increases in quadriceps motor cortex excitability and improved cycling performance^44,48–50^, suggesting a vertex–shoulder montage can effectively modulate lower extremity motor areas while avoiding cortical inhibition at the cathode.

For the present trial, we modelled a Cz–shoulder montage using SIMNIBS Ernie Extended^51^, with 5 cm electrodes and default tissue conductivities (Figure 2). Electrical field modelling indicated robust fields in both the leg motor area (maximal 0.80, mean 0.46 volts/m) and cerebellum (maximal 1.24, mean 0.86), supporting the Cz–shoulder configuration as a biologically plausible approach for targeting lower extremity motor cortex and cerebellar pain networks in SCI-NP. It is also important to note that this montage would direct current closer to the EEG-NF electrode sites (C1 and C2), which may be preferable to a standard montage in which current enters predominantly through one side of the motor cortex (e.g., C3) but not the other.

**Figure 2.**
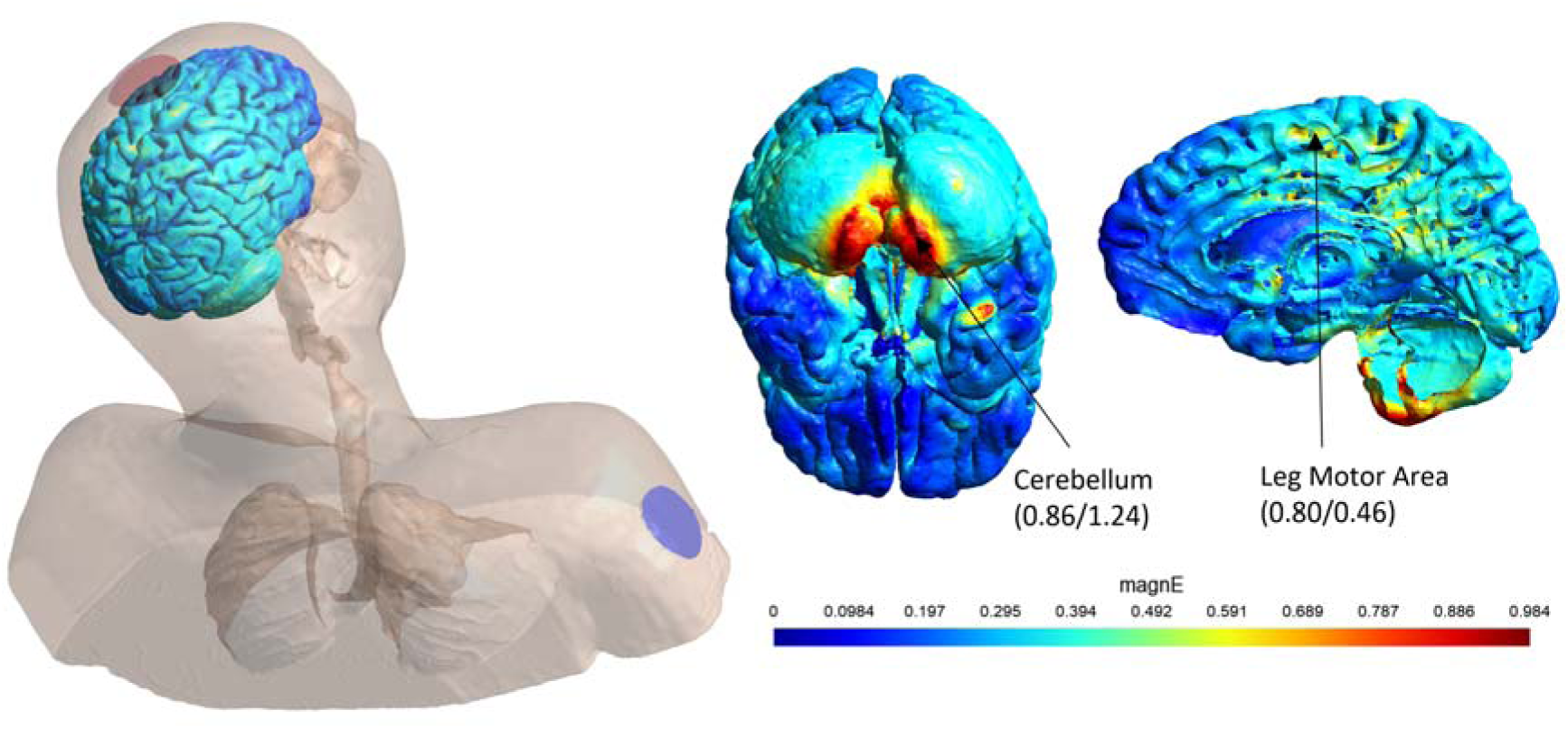
Results of Electrical field modelling using a novel Montage with the anode positioned on Cz and return electrode placed on the right shoulder. E fields are generated in both the leg motor area and cerebellum.

In this trial, the anode at the vertex will be secured with a 3D-printed headset (Figure 3) and the shoulder cathode with a band, both designed for home use. Detailed instructions are provided through printed manuals, step-by-step guidance in the tablet app, and a supervised initial online session.

**Figure 3.**
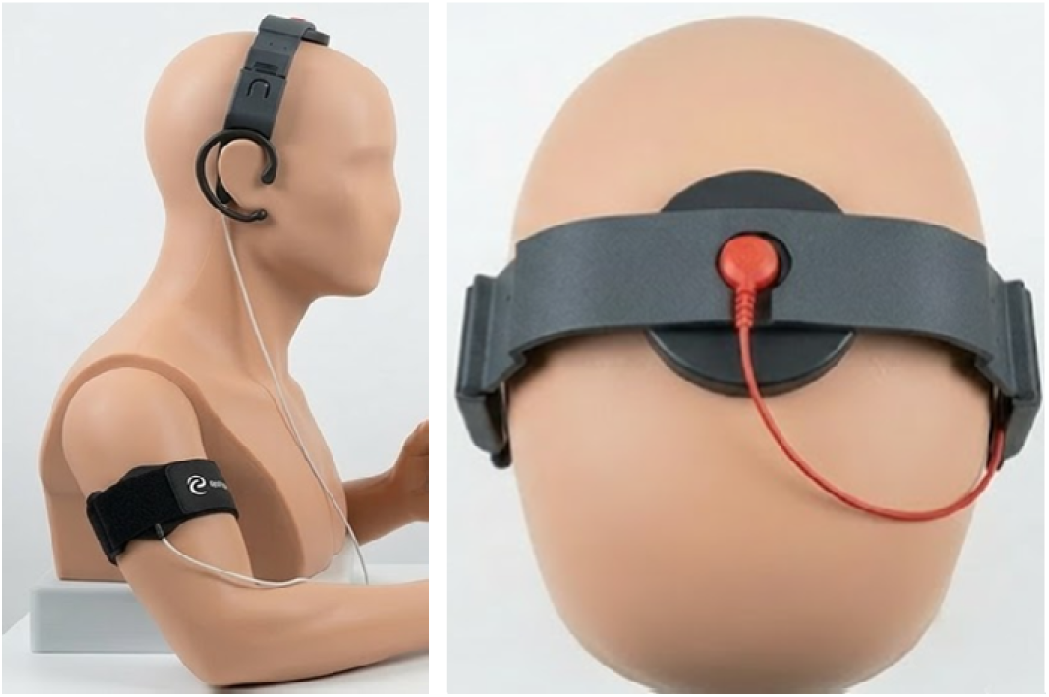
The building blocks of the montage, including a 3D printed headset that will contain the anodal electrode and armband which will contain the cathodal electrode.

### Time window between tDCS and EEG-NF (Groups 1 and 2 only)

Once the tDCS is complete, participants will be instructed to clean residual tDCS saline (comb out crystals, rinse and wipe with water, gauze, cotton swabs, then dry with gauze and alcohol wipes). tDCS is thought to prime the brain by increasing cortical excitability and enhancing plasticity in neural circuits engaged by concurrent therapies^52^. However, our system cannot deliver EEG-NF and tDCS simultaneously because tDCS introduces electrical artefacts that disrupt the neurofeedback algorithm, so EEG-NF must follow tDCS. To determine how long motor cortical excitability remains elevated after 2mA anodal motor cortex tDCS, we collected pilot data from 20 individuals with an SCI (13 of whom had neuropathic pain), measuring motor evoked potentials (MEPs) every 5 minutes for 60 minutes. Figure 4 shows the mean MEP amplitude at each timepoint, normalised to the pre-tDCS MEP amplitude; error bars indicate 95% confidence intervals. The confidence intervals remained consistently above the pre-tDCS reference (i.e., 1.0) until at least 45 minutes post-tDCS, suggesting a sustained period of heightened corticomotor excitability. To ensure EEG-NF is administered within this window, we will commence approximately 10 minutes after tDCS, with this interval used for EEG-NF setup.

**Figure 4.**
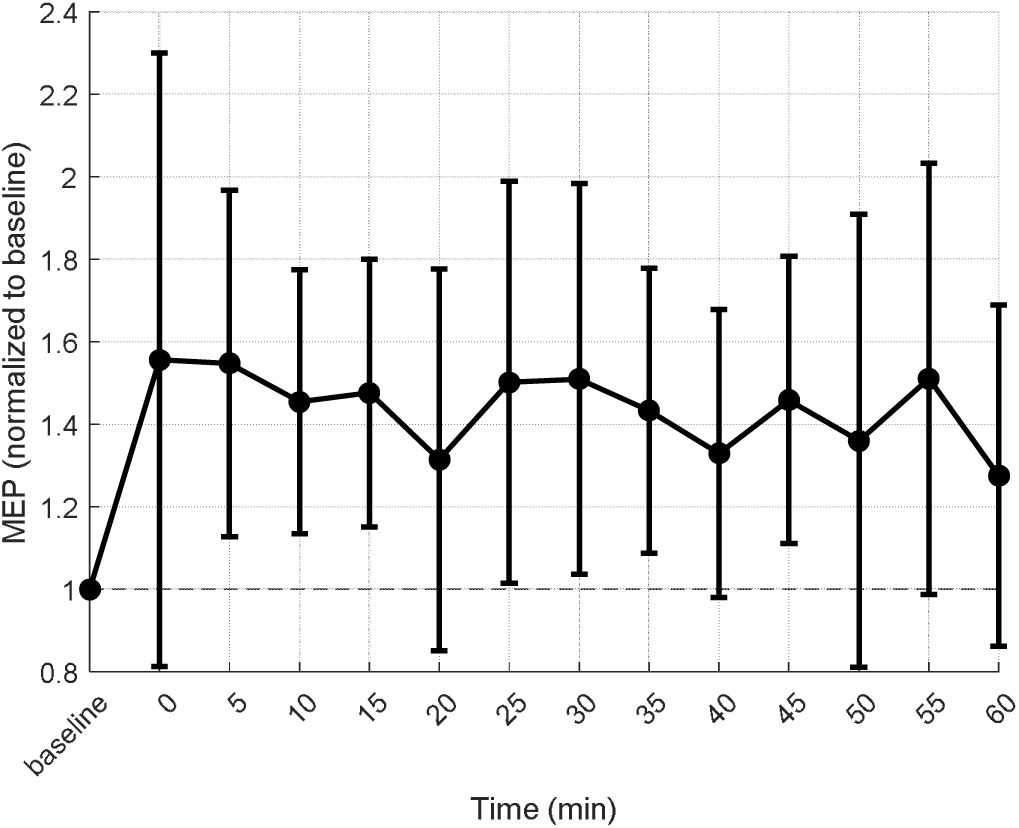
Changes in corticomotor excitability within a 60-minute period following 10 minutes of 2mA anodal tDCS to the left primary motor cortex in 20 individuals with a SCI. The error bars represent the 95% confidence interval.

### EEG Neurofeedback Intervention (Groups 1 and 2 Only)

A wireless EEG headset was custom-designed for this trial using a variant of the OpenBCI Ganglion board (openbci.com). Several 3D-printed prototypes were iteratively tested in several able-bodied participants to optimise comfort, usability, and signal quality ^53^. The final one-size-fits-all design includes adjustable sliding earpieces and two saline-soaked sponge electrodes at C1 and C2 (10–20 system) targeting sensorimotor cortex, with two ear-clip reference electrodes.

A tablet app guides participants through headset setup, impedance checks, resting-state EEG, and neurofeedback (Figure 5). Before each session, switch on the headset, connect via Bluetooth, insert pre-soaked electrodes at C1/C2, and attach ear-clips. Impedance is checked automatically; if >50 kΩ, on-screen troubleshooting is provided.

**Figure 5.**
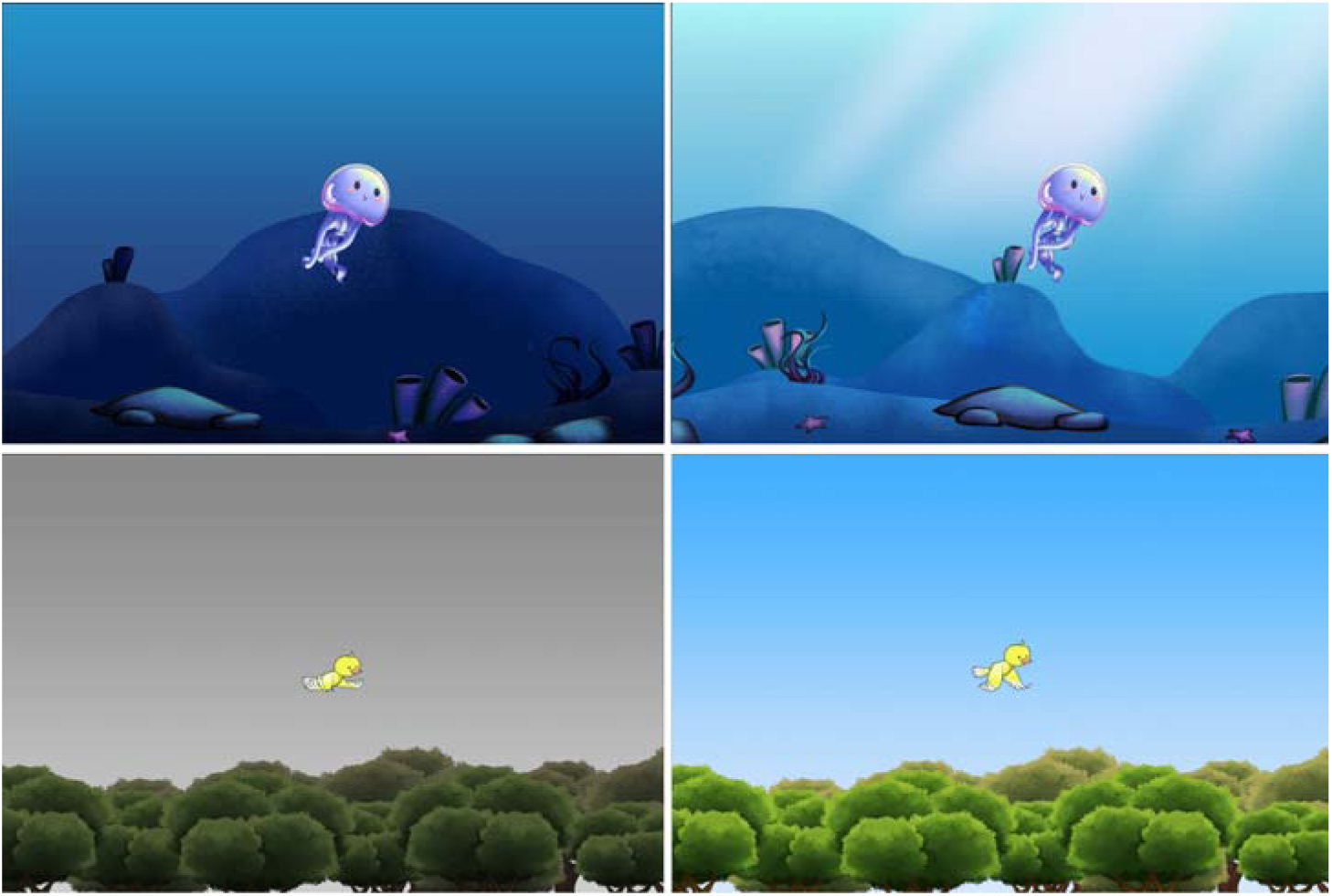
Jellyfish Game: The jellyfish swims through the bright blue ocean as participants successfully regulate the targeted frequency bands. B. Bird game: The Bird flies through the blue sky in response to participants’ regulation of the targeted frequency bands.

Each session will start with 2 minutes of eyes-open resting-state EEG to establish individual alpha, theta, and beta baselines. Participants will then complete five 2.5-minute neurofeedback game rounds, with short breaks. Four games (Jellyfish, Bird, Plane, Rocket) will be introduced sequentially to maintain engagement. The protocol trains suppression of theta (4–7 Hz) and high-beta (20–30 Hz), and enhancement of sensorimotor rhythms (SMR, 10–15 Hz, constituting high alpha and low beta). When all three bands move in the desired direction, the games provide real-time visual feedback via character motion and background changes; scores at the end of each round track progress. Participants will be encouraged to develop their own mental strategies (e.g., positive thinking, focus on game elements, pleasant imagery, breathing techniques). Relaxing background music can be adjusted or muted. Post-intervention interviews will explore strategies perceived as most effective.

### Summary of the Interventions

A summary of each intervention session is provided in Figure 6.

**Figure 6.**
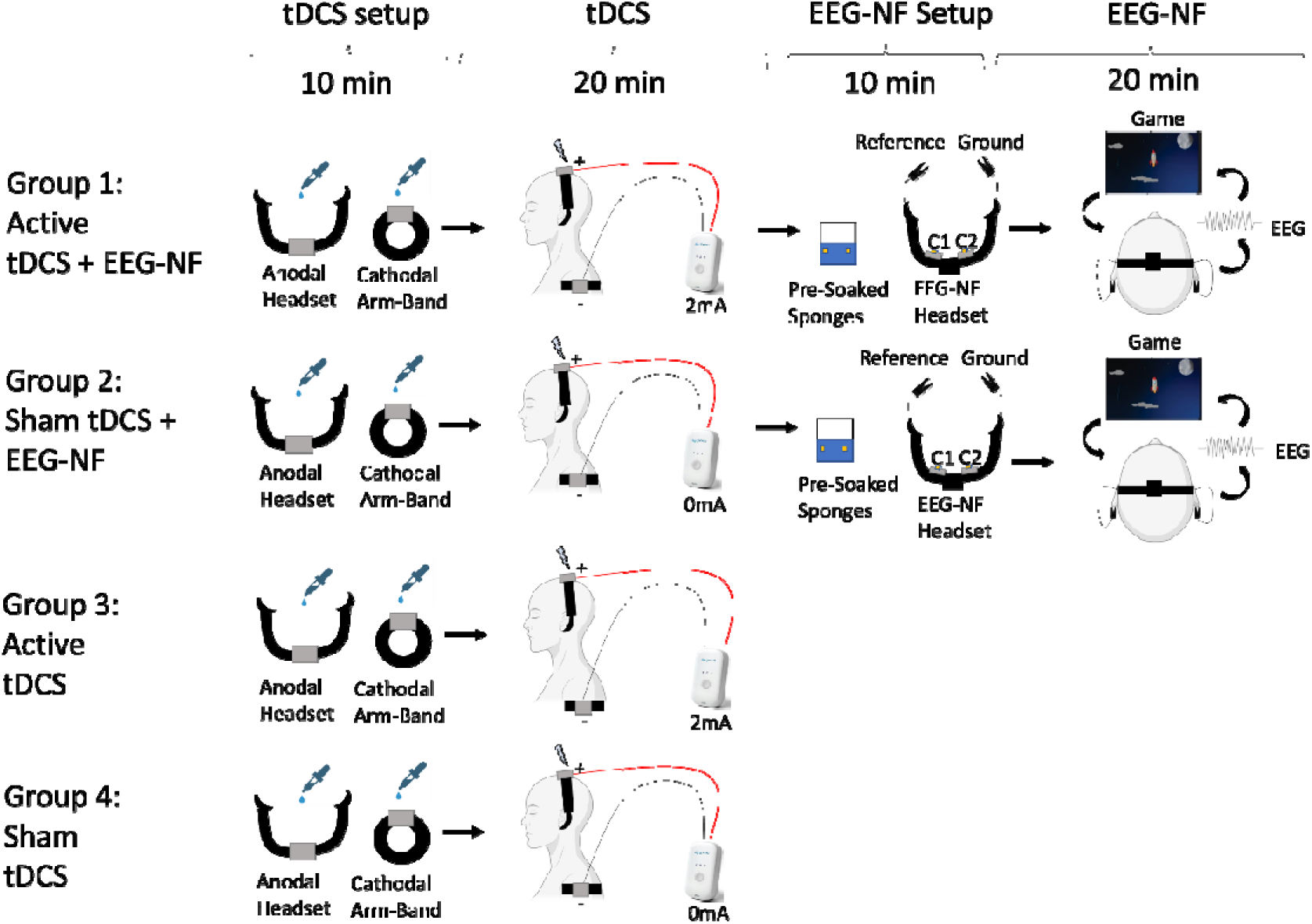
Overview of the Intervention in each of the 4 groups. All groups will begin with the setup of transcranial direct current stimulation (tDCS), which takes approximately 10 minutes. This involves placing a headset and a shoulder armband, which respectively contain the anode (positioned over Cz) and the cathode (positioned over the shoulder). Further justification for this electrode montage is provided in the main text. Both electrodes are prepared by applying saline to a foam pad located within the electrode casing. Once the setup is complete and impedance levels are confirmed to be acceptable, participants will undergo a 20-minute tDCS session. During stimulation, participants will remain seated and relaxed. Participants in Groups 1 and 3 will receive active tDCS (i.e., 2mA constant current), while those in Groups 2 and 4 will receive sham tDCS (i.e., a brief ramp-up to 2 mA followed by a constant current of 0 mA to mimic the sensation of stimulation without the neuromodulatory effects). Following tDCS, participants in Groups 1 and 2 will proceed to setup for the EEG-NF session, which also takes approximately 10 minutes. This setup involves placing pre-soaked sponges over electrodes C1 and C2 of the EEG-NF headset. Once impedance levels are confirmed to be below 50kΩ, participants will begin a 20-minute EEG-NF session.

### Outcome Measures

Table 2 shows the schedule of enrolment, interventions and assessments. Responses to all outcome measures will be self-entered and recorded online directly through REDCap^54^. At baseline, an online questionnaire will be sent to participants requesting demographics (age, birth sex, gender identity, marital status, education, employment, household income, and country of birth) and injury information (time since injury, injury level, aetiology of injury).

**Table 2.** Schedule of enrolment, interventions and assessments.

| Timepoint | Enrolment | Baseline | Randomisation | 20 Sessions within 5 weeks | 6 weeks | 16 weeks | 26 weeks | 52 weeks |
| --- | --- | --- | --- | --- | --- | --- | --- | --- |
| **PRE-INTERVENTION** |  |  |  |  |  |  |  |  |
| Eligibility Screening | X |  |  |  |  |  |  |  |
| Informed Consent | X |  |  |  |  |  |  |  |
| Baseline Questionnaires |  | X |  |  |  |  |  |  |
| Randomisation Module |  |  | X |  |  |  |  |  |
| **INTERVENTIONS:** |  |  |  |  |  |  |  |  |
| Active tDCS |  |  |  | X |  |  |  |  |
| Sham tDCS |  |  |  | X |  |  |  |  |
| EEG-NF + Active tDCS |  |  |  | X |  |  |  |  |
| EEG-NF + Sham tDCS |  |  |  | X |  |  |  |  |
| **ASSESSMENTS:** |  |  |  |  |  |  |  |  |
| BPI-Pain Severity |  | X |  |  | X | X | X | X |
| BPI-Pain Interference |  | X |  |  | X | X | X | X |
| BPI-Worst Pain |  | X |  |  | X | X | X | X |
| Sleep assessment |  | X |  |  | X | X | X | X |
| PHQ-9 |  | X |  |  | X | X | X | X |
| SF-36 |  | X |  |  | X | X | X | X |

#### Primary Outcome: Pain Severity

The primary outcome is the pain severity assessed 6 weeks post-randomisation (primary endpoint). Pain severity will be assessed using the Brief Pain Inventory (BPI) - Pain Severity scale. Participants will be asked to rate their worst and least over the last 7 days in addition to their average and current pain, on a scale from 0 (no pain) to 10 (worst imaginable pain). Pain severity will be operationalised as the average of these four items.

#### Secondary Outcomes

- **Pain Severity**: BPI Pain Severity assessed at 16 weeks, 26 weeks, and 52 weeks post-randomisation Other secondary outcomes (listed below) will be measured at baseline, 6 weeks, 16 weeks, 26 weeks, and 52 weeks post-randomisation.
- **Worst Pain: ‘**Pain at its worst’ item from BPI-Pain Severity
- **Pain Interference:** BPI Pain Interference, modified for SCI
- **Sleep Disturbance:** PROMIS Sleep Disturbance 8b Short Form + 2 items of Medical Outcomes Study Sleep Scale (MOS-SS)
- **Mood:** Patient Health Questionnaire (PHQ-9)
- **Health-related quality of life:** Short-Form Survey (SF-36), modified for SCI

### Statistical Analyses

A detailed, date-stamped Statistical Analysis Plan (SAP) will be uploaded to the Open Science Framework. Key elements are summarised here.

#### General principles

Analyses will emphasise estimation of causal effects with point estimates and confidence intervals, supplemented by two-sided tests^55,56^. A blinded “dummy” analysis (artificial re-randomisation) will be used to finalise modelling choices. Reporting will follow CONSORT (including non-pharmacological/sham extensions) ^57^. All randomised participants will be analysed under an intention-to-treat framework.

#### Estimands

Primary estimands are the average causal effects of (1) the effect of EEG-NF compared to no EEG-NF when given sham tDCS, (2) the effect of tDCS compared to sham tDCS when not given EEG-NF, and (3) the effect of combined tDCS + EEG-NF together compared to no intervention (sham tDCS), on BPI–Pain Severity at 6 weeks.

#### Multiplicit

Each of the three primary objectives (EEG-NF, tDCS, combined) will use α = 0.05 (two-sided) without formal multiplicity adjustment, consistent with a “three-trials-in-one” design.

#### Primary analysis of effectiveness

For the primary endpoint (pain severity at 6 weeks), the primary analysis will use an optimally efficient factorial regression model with BPI–Pain Severity as the dependent variable, baseline BPI–Pain Severity as a covariate, and fixed indicators for EEG-NF (yes/no), tDCS (active/sham), and their interaction (EEG-NF×tDCS). This model provides estimates of each intervention’s effect and their combination, adjusted for baseline values. From this model, three pre-specified pairwise comparisons will be estimated using linear contrasts: (1) EEG-NF vs no EEG-NF within sham tDCS, (2) active vs sham tDCS within no EEG-NF, and (3) combined EEG-NF + active tDCS vs sham tDCS. These contrasts will be estimated within a single factorial model, enabling interpretation of each intervention’s effect conditional on the absence of the other. Results will be reported as mean differences (negative favouring active), with 95% confidence intervals and two-sided p-values, focusing on estimation.

#### Missing data

Primary analyses will use all available data assuming missing at random^58^. We will report missingness by arm/timepoint. If 5–40% of 6-week outcomes are missing, a sensitivity analysis using multiple imputation (Bayesian MCMC under a multivariate normal model; 50 imputations; Rubin’s rules) will be conducted ^59^:

### Ethics and Dissemination

The protocol has been approved by the University of New South Wales Human Research Ethics Committee (iRECS9099). Any protocol deviations will require ethics amendments and registry updates. Identifying information will be replaced with unique IDs, and personal data will be kept confidential in line with NHMRC guidelines and stored in a secure, password-protected database accessible only to the research team for 15 years. Findings will be disseminated via peer-reviewed publications and conference presentations.

### Trial Status

The trial is scheduled to begin in June 2026 and end in December 2029.

## Discussion

This trial addresses a major unmet need for addressing neuropathic pain after SCI, a common, highly disabling secondary condition, which is often refractory to existing pharmacological and non-pharmacological treatments^1–6^. By targeting thalamocortical dysrhythmia and aberrant sensorimotor rhythms using EEG-NF and cortical excitability with a mechanistically informed tDCS montage, the trial directly tests promising, low-risk neuromodulatory approaches that can be delivered at home, reducing access barriers for people with significant mobility limitations. The trial is powered based on an established MCID for pain and incorporates long-term follow-up to assess durability of benefit. Rigorous methods, including prospective registration, SPIRIT- and CONSORT-consistent reporting, pre-specified estimands and a detailed SAP are designed to maximise robustness and interpretability. If EEG-NF and/or tDCS prove effective, alone or in combination, the findings will provide a strong evidential and mechanistic basis for integrating home-based neuromodulatory therapies into routine care pathways for SCI-NP.

## Data Availability

This is a trial protocol with no new data

## Conflict of Interest

The authors declare that they have no competing interests

## Funding

This work was supported by the Medical Research Future Fund awarded to Sylvia M Gustin

## Notes

### Competing Interest Statement

The authors have declared no competing interest.

### Clinical Trial

ACTRN12625000737437

### Author Declarations

The protocol has been approved by the University of New South Wales Human Research Ethics Committee (iRECS9099)

## References

1. Bresnahan, J.J., Scoblionko, B.R., Zorn, D., Graves, D.E. & Viscusi, E.R. The demographics of pain after spinal cord injury: a survey of our model system. Spinal cord series and cases 8, 14 (2022).

2. Boldt, I., et al. Non-pharmacological interventions for chronic pain in people with spinal cord injury. Cochrane Database of Systematic Reviews (2014).

3. Siddall, P.J. Management of neuropathic pain following spinal cord injury: now and in the future. Spinal cord 47, 352–359 (2009).

4. Rosner, J., et al. Central neuropathic pain. Nature Reviews Disease Primers 9, 73 (2023).

5. Fakhri, S., Abbaszadeh, F. & Jorjani, M. On the therapeutic targets and pharmacological treatments for pain relief following spinal cord injury: A mechanistic review. Biomedicine & Pharmacotherapy 139, 111563 (2021).

6. Eller, O.C., Willits, A.B., Young, E.E. & Baumbauer, K.M. Pharmacological and non-pharmacological therapeutic interventions for the treatment of spinal cord injury-induced pain. Frontiers in Pain Research 3, 991736 (2022).

7. Gustin, S., et al. Thalamic activity and biochemical changes in individuals with neuropathic pain after spinal cord injury. PAIN® 155, 1027–1036 (2014).

8. Jensen, M., et al. Brain EEG activity correlates of chronic pain in persons with spinal cord injury: clinical implications. Spinal Cord 51, 55–58 (2013).

9. Boord, P., et al. Electroencephalographic slowing and reduced reactivity in neuropathic pain following spinal cord injury. Spinal cord 46, 118–123 (2008).

10. Mussigmann, T., Bardel, B. & Lefaucheur, J.-P. Resting-state electroencephalography (EEG) biomarkers of chronic neuropathic pain. A systematic review. Neuroimage 258, 119351 (2022).

11. Jensen, M.P., et al. Baseline brain activity predicts response to neuromodulatory pain treatment. Pain Medicine 15, 2055–2063 (2014).

12. Vučković, A., Altaleb, M.K.H., Fraser, M., McGeady, C. & Purcell, M. EEG correlates of self-managed neurofeedback treatment of central neuropathic pain in chronic spinal cord injury. Frontiers in Neuroscience 13, 762 (2019).

13. Ibric, V.L. & Dragomirescu, L.G. Neurofeedback in pain management. Introduction to quantitative EEG and neurofeedback: Advanced theory and applications 2nd ed, 417–451 (2009).

14. Miró, J., et al. Psychological neuromodulatory treatments for young people with chronic pain. Children 3, 41 (2016).

15. Micoulaud-Franchi, J.-A., et al. Electroencephalographic neurofeedback: Level of evidence in mental and brain disorders and suggestions for good clinical practice. Neurophysiologie Clinique/Clinical Neurophysiology 45, 423–433 (2015).

16. Hershaw, J.N., Hill-Pearson, C.A., Arango, J.I., Souvignier, A.R. & Pazdan, R.M. Semi-automated neurofeedback therapy for persistent postconcussive symptoms in a military clinical setting: a feasibility study. Military medicine 185, e457–e465 (2020).

17. Caro, X.J. & Winter, E.F. EEG biofeedback treatment improves certain attention and somatic symptoms in fibromyalgia: a pilot study. Applied psychophysiology and biofeedback 36, 193–200 (2011).

18. Rice, D.A., et al. Home-based EEG Neurofeedback for the Treatment of Chronic Pain: A Randomized Controlled Clinical Trial. The Journal of Pain 25, 104651 (2024).

19. Sorger, B., Scharnowski, F., Linden, D.E., Hampson, M. & Young, K.D. Control freaks: Towards optimal selection of control conditions for fMRI neurofeedback studies. Neuroimage 186, 256–265 (2019).

20. Lütolf, R., Rosner, J., Curt, A. & Hubli, M. Indicators of central sensitization in chronic neuropathic pain after spinal cord injury. European Journal of Pain 26, 2162–2175 (2022).

21. Nitsche, M.A. & Paulus, W. Excitability changes induced in the human motor cortex by weak transcranial direct current stimulation. The Journal of physiology 527, 633 (2000).

22. Pacheco-Barrios, K., et al. Methods and strategies of tDCS for the treatment of pain: current status and future directions. Expert review of medical devices 17, 879–898 (2020).

23. O’Connell, N.E., Marston, L., Spencer, S., DeSouza, L.H. & Wand, B.M. Non-invasive brain stimulation techniques for chronic pain. Cochrane database of systematic reviews (2018).

24. Soliman, N., et al. Pharmacotherapy and non-invasive neuromodulation for neuropathic pain: a systematic review and meta-analysis. The Lancet Neurology 24, 413–428 (2025).

25. Chowdhury, N.S., Bikson, M. & Gustin, S. A roadmap for transcranial direct current stimulation for chronic pain. PAIN® In Press(2025).

26. Li, L., et al. Non-invasive brain stimulation for neuropathic pain after spinal cord injury: a systematic review and network meta-analysis. Frontiers in Neuroscience 15, 800560 (2022).

27. Moreno-Duarte, I., et al. Targeted therapies using electrical and magnetic neural stimulation for the treatment of chronic pain in spinal cord injury. Neuroimage 85, 1003–1013 (2014).

28. Lefaucheur, J.-P. & Wendling, F. Mechanisms of action of tDCS: A brief and practical overview. Neurophysiologie Clinique 49, 269–275 (2019).

29. Soler, M.D., et al. Effectiveness of transcranial direct current stimulation and visual illusion on neuropathic pain in spinal cord injury. Brain 133, 2565–2577 (2010).

30. Soler, D., Moriña, D., Kumru, H., Vidal, J. & Navarro, X. Transcranial direct current stimulation and visual illusion effect according to sensory phenotypes in patients with spinal cord injury and neuropathic pain. The Journal of Pain 22, 86–96 (2021).

31. Chan, A.-W., et al. SPIRIT 2013 explanation and elaboration: guidance for protocols of clinical trials. Bmj 346(2013).

32. Turner, L., et al. Consolidated standards of reporting trials (CONSORT) and the completeness of reporting of randomised controlled trials (RCTs) published in medical journals. Cochrane database of systematic reviews (2012).

33. Busse, J.W., et al. Optimal strategies for reporting pain in clinical trials and systematic reviews: recommendations from an OMERACT 12 workshop. The Journal of rheumatology 42, 1962–1970 (2015).

34. Gustin, S.M., et al. Cortical mechanisms underlying immersive interactive virtual walking treatment for amelioration of neuropathic pain after spinal cord injury: Findings from a preliminary investigation of thalamic inhibitory function. Journal of Clinical Medicine 12, 5743 (2023).

35. Bryce, T., et al. Screening for neuropathic pain after spinal cord injury with the spinal cord injury pain instrument (SCIPI): a preliminary validation study. Spinal Cord 52, 407–412 (2014).

36. Mehta, S., McIntyre, A., Guy, S., Teasell, R.W. & Loh, E. Effectiveness of transcranial direct current stimulation for the management of neuropathic pain after spinal cord injury: a meta-analysis. Spinal Cord 53, 780–785 (2015).

37. Chowdhury, N.S., et al. Predicting individual pain sensitivity using a novel cortical biomarker signature. JAMA neurology (2025).

38. Liu, X., et al. Corticospinal Excitability in Bilateral M1 Hand Areas: Association with Neuropathic Pain After Spinal Cord Injury. J Pain Res 18, 4003–4018 (2025).

39. Gangitano, M., et al. Modulation of input–output curves by low and high frequency repetitive transcranial magnetic stimulation of the motor cortex. Clinical Neurophysiology 113, 1249–1257 (2002).

40. Ciampi de Andrade, D. & García-Larrea, L. Beyond trial-and-error: Individualizing therapeutic transcranial neuromodulation for chronic pain. European Journal of Pain 27, 1065–1083 (2023).

41. Floyd, J.T., et al. Transcranial Direct Current Stimulation (tDCS) can alter cortical excitability of the lower extremity in healthy participants: A review and methodological study. Frontiers in neurology and neuroscience research 1, 100002 (2020).

42. Kaski, D., Quadir, S., Patel, M., Yousif, N. & Bronstein, A.M. Enhanced locomotor adaptation aftereffect in the “broken escalator” phenomenon using anodal tDCS. Journal of neurophysiology 107, 2493–2505 (2012).

43. Tomio, R., Akiyama, T., Ohira, T. & Yoshida, K. Effects of transcranial stimulating electrode montages over the head for lower-extremity transcranial motor evoked potential monitoring. Journal of neurosurgery 126, 1951–1958 (2016).

44. Angius, L., Hopker, J.G., Marcora, S.M. & Mauger, A.R. The effect of transcranial direct current stimulation of the motor cortex on exercise-induced pain. European journal of applied physiology 115, 2311–2319 (2015).

45. Kong, Q., Li, T., Reddy, S., Hodges, S. & Kong, J. Brain stimulation targets for chronic pain: Insights from meta-analysis, functional connectivity and literature review. Neurotherapeutics 21, e00297 (2024).

46. Bocci, T., et al. Cerebellar transcranial direct current stimulation (ctDCS) ameliorates phantom limb pain and non-painful phantom limb sensations. The Cerebellum 18, 527–535 (2019).

47. Bocci, T., et al. Cerebellar direct current stimulation modulates pain perception in humans. Restorative Neurology and Neuroscience 33, 597–609 (2015).

48. Seamon, B.A., Bowden, M.G., Kindred, J.H., Embry, A.E. & Kautz, S.A. Transcranial direct current stimulation electrode montages may differentially impact variables of walking performance in individuals poststroke: a preliminary study. Journal of Clinical Neurophysiology 40, 71–78 (2023).

49. Tatemoto, T., Yamaguchi, T., Otaka, Y., Kondo, K. & Tanaka, S. Anodal transcranial direct current stimulation over the lower limb motor cortex increases the cortical excitability with extracephalic reference electrodes. in Converging Clinical and Engineering Research on Neurorehabilitation 829–834 (Springer, 2013).

50. Noetscher, G.M., Yanamadala, J., Makarov, S.N. & Pascual-Leone, A. Comparison of cephalic and extracephalic montages for transcranial direct current stimulation—a numerical study. IEEE Transactions on Biomedical Engineering 61, 2488–2498 (2014).

51. Van Hoornweder, S., et al. Head and shoulders—The impact of an extended head model on the simulation and optimization of transcranial electric stimulation. Imaging Neuroscience 2, 1–11 (2024).

52. Hurley, R. & Machado, L. Using tDCS priming to improve brain function: Can metaplasticity provide the key to boosting outcomes? Neuroscience & Biobehavioral Reviews 83, 155–159 (2017).

53. Hesam-Shariati, N., et al. A home-based self-directed EEG neurofeedback intervention for people with chronic neuropathic pain following spinal cord injury (the StoPain Trial): description of the intervention. Spinal Cord 62, 658–666 (2024).

54. Harris, P.A., et al. The REDCap consortium: building an international community of software platform partners. Journal of biomedical informatics 95, 103208 (2019).

55. Gardner, M.J. & Altman, D.G. Confidence intervals rather than P values: estimation rather than hypothesis testing. Br Med J (Clin Res Ed) 292, 746–750 (1986).

56. Amrhein, V., Greenland, S. & McShane, B. Scientists rise up against statistical significance. Nature 567, 305–307 (2019).

57. Boutron, I., et al. Extending the CONSORT statement to randomized trials of nonpharmacologic treatment: explanation and elaboration. Annals of internal medicine 148, 295–309 (2008).

58. Fitzmaurice, G.M., Laird, N.M. & Ware, J.H. Applied longitudinal analysis, (John Wiley & Sons, 2012).

59. Jakobsen, J.C., Gluud, C., Wetterslev, J. & Winkel, P. When and how should multiple imputation be used for handling missing data in randomised clinical trials–a practical guide with flowcharts. BMC medical research methodology 17, 162 (2017).

